# The impact of changes in mental health services in response to COVID-19 on people with mental health conditions: protocol for a rapid review

**DOI:** 10.1101/2022.02.15.22270931

**Authors:** G Yu, D Craig, Y Fu

## Abstract

**Introduction:** The COVID-19 pandemic has caused disruptions to mental health services, forcing the rapid implementation of alternative ways of delivering services alongside a greater immediate, and continuously growing, demand across those services. The care and level of mental health service provided are felt to be inadequate to respond to the increasing demand for mental health conditions in the time of the pandemic, leading to an urgent need to learn from service change and consequences to inform solutions and plans to support the NHS post-pandemic plan in the UK. This rapid review aims to understand the changes in mental health services during the pandemic and summarise the impact of these changes on the health outcomes of people with mental health conditions.

**Methods and analysis:** Cochrane CENTRAL, MEDLINE, EMBASE, and PsycInfowill be searched for eligible studies with key terms indicating mental health AND COVID-19 AND health services. Studies will be included if objective and subjective effects of changes to mental health services in response to COVID-19 are reported on adults with mental health conditions, peer-reviewed and published in the English language. Study selection and data extraction will be undertaken dependently by two reviewers. Evidence will be summarised narratively and in a logic model.

**Ethics and dissemination:** Ethics approval is not required for this review. A list of interventions/services/models of care delivered to people with mental health conditions will be grouped as “Do”, “Don’t” and “Don’t know” based on the evidence on effectiveness and acceptability. The results will be written for publication in an open-access peer-reviewed journal and disseminated to the public and patients, clinicians, commissioners, funders, and academic conferences.

**PROSPERO registration number:** CRD42022306923

**Strengths and limitations of this study:**

- This is a rapid review with a systematic search of literature
- Evidence on changes to mental health services and associated health outcomes will be examined to make practice recommendations
- This review will provide the evidence base to inform solutions and plans to support the mental health service provision for the post-pandemic period
- Some limitations to the study design include limited to OECD studies, exclusion of non-English studies, publication bias, quality of data, selection bias, and no quality assessment in the rapid evidence review.

## INTRODUCTION

In the UK, up to December 2021, 1 in 7 people have had a positive COVID-19 test result. It is recognised that as well as the physical effects, a COVID-19 diagnosis has adverse mental health effects both in previously healthy people and those with pre-existing mental health conditions^1^. About 1 in 5 people with COVID-19 have experienced poorer mental health within 90 days of diagnosis^2^. It is estimated that the pandemic will lead to new or additional mental health support for up to 10 million people in England (around 20% of the population).^3^ Evidence from previous studies illustrated that the virus and periods of lockdown have deteriorated population mental health and disproportionately worsened the mental health burden for certain population subgroups including those with pre-existing mental health conditions.^4 5^ People with existing psychiatric diagnoses have reported increased symptoms and poorer access to services and support leading to relapse and suicidal behaviour^1^. At the same time, they are more likely to contract COVID-19 than people with no history of poor mental health. The adaptations required to enable the delivery of mental healthcare during this period of extended infection-control measures could have been disproportionally detrimental to those now living with mental health conditions that have arisen since the first UK-wide lockdown began in March 2020. Difficulties attending review appointments in person and closure of support services are likely to have impacted all those in, or in need of, active treatment.^6^ The unequal impact of the pandemic is likely to further entrench and exacerbate the existing structural inequalities in mental health among people with pre-existing mental health conditions before COVID-19, and the mental health services provided have failed to meet their increasing demand for mental health conditions during the time of the COVID-19 pandemic.^7^

The NHS has set up a long term plan to improve mental healthcare services that are widely regarded as being under-resourced.^8^ However, for people with mental health conditions, there is an incomplete picture of the impact of the pandemic on the pattern of mental health services. Despite bringing current service inadequacies to the forefront, the pandemic could provide an opportunity to rethink conventional approaches to mental health services planning to meet patients’ needs. For example, remote community treatment and support has long been suggested but has not previously been implemented widely because of barriers and challenges from both healthcare staff and service users. Since the onset of the pandemic, the situation has changed.^9^ Similarly, the threshold for hospital admission for mental illness varies between individuals and requires continuous adaptation over time. Therefore, learning from health service changes throughout the pandemic, and their consequences for people’s physical and mental health is vital to inform practical policy solutions for integrated service recovery and effectively plan services that reach those with the greatest need.

The WHO recommends rapid review methods as an efficient approach to provide rapid but relevant and contextualised evidence to the health decision-makers when there are time, resources or other logistical constraints.^10-15^ We will conduct a rapid review to synthesise the evidence on changes in mental health services because of the COVID-19 pandemic and their impacts in high-income countries.^16 17^

## METHODS AND ANALYSIS

A rapid review will be undertaken to provide an evidence base supporting the recommendation of mental health services and identify areas where the evidence base is lacking, and future research is required. The review will be guided by the Cochrane guidance for rapid reviews ^18^. Preferred Reporting Items for Systematic reviews and Meta-Analyses extension for Rapid Reviews guidance^19^ will be followed for reporting.

This protocol has been developed in advance of the review to improve the transparency and quality of the methods to help reduce bias and enhance the reproducibility of the results. This has been registered with the PROSPECT CRD42022306923.

### Eligibility

#### Type of studies

Peer-reviewed quantitative or qualitative empirical studies describing the setting, problems addressed, resource requirements, aim, service components, provider, method of delivery, objective and subjective effects of changes to mental health services in response to COVID-19 will be included. Included studies will be those undertaken in an Organisation for Economic Co-operation and Development (OECD) country^20^, to ensure a degree of commonality in the health system and socioeconomic and demographic context.

#### Type of participants

People aged 18 or over experiencing mental health conditions as described by NHS^21^ who were in need of mental health support during the pandemic.

#### Type of health services

Interventions, services and models of care delivered in response to COVID-19 to provide support for adults with mental health conditions will be included.

#### Type of outcome measures

Primary outcomes are objective measures and subjective effects of changes, efficacy or use of a service by mental health patients. Secondary outcomes are changes in knowledge, attitudes or satisfaction of service users and/or professionals and health inequalities.

### Search methods

Cochrane CENTRAL, MEDLINE, Embase and PsycInfowill be searched for from 2019 to the present. A search strategy has been developed for MEDLINE with support from an independent information specialist, using a range of keywords and subject headings representing COVID-19, mental health and low- and middle-income countries (see Appendix). This will be used to inform the detailed search strategy for other databases. Reference lists and citation indexes of relevant studies will also be examined. Only OECD studies published in or after 2019 and in the English language (no resource available for translation) will be searched.

### Selection of studies

Studies identified from databases will be exported to EndNote X9^22^ for deduplication. Study titles of abstracts will be screened independently according to the selection criteria. Any results that are inconclusive at the initial screen will be included and considered at full-text screening. All full-text papers will be screened independently by two researchers (GY and YF). Any discrepancies will be resolved by discussion and consensus. Where there is a disagreement between two reviewers, a third researcher (DC) will be consulted to reach a consensus.

### Data extraction

A data extraction sheet will be designed to capture information including author’s first name, publication date, setting, study design, sample size, mental health conditions, characteristics of participants, service components, service provider, method of delivery, resources required, outcome measures and main study results. GY will extract all the data. YF will check for accuracy and completeness through random double-extraction of 10% of included studies. Where a study appears to have multiple citations, original authors will be contacted for clarification. All information from multiple citations will be used if no replies are received.

### Quality assessment and quality control

This is a rapid review, so no quality assessment will be conducted. This can be done retrospectively if time and resources allow.

The following steps will be taken to ensure quality control for the searching, screening, data extraction, and coding process. GY will conduct screening and data extraction following pre-determined inclusion criteria and data extraction framework. For articles that are retrieved and full text saved, YF will check 10% of the coding to ensure they meet the screening criteria. Where there is a disagreement between two reviewers, a third researcher (DC) will be consulted to reach a consensus. Synthesis of each outcome will be conducted by GY and independently revised by YF.

### Data synthesis and analysis

A tabulated and narrative synthesis of the results will be undertaken following current best practice^23-25^ to conduct synthesis systematically and transparently. It will focus on the mental health services, mechanisms and their impact on health outcomes. A logic model will be produced to present context, service provision and outcomes. Possible unintended adverse outcomes will also be reported. Also, a list of interventions/services/models of care delivered to people with mental health conditions will be grouped as “Do”, “Don’t” and “Don’t know” based on the strength of the evidence on effectiveness and acceptability.

If data is available, outcomes of studies will be synthesised according to characteristics of study participants, for example, deprived communities, ethnic minorities, to produce evidence on health inequalities that is likely to have been exacerbated during the pandemic.

## PATIENT AND PUBLIC INVOLVEMENT

This study has been designed and developed in consultation with two public members (one with lived experience), to ensure their input on the study design. They both read and commented on the review summary, search strategies, eligibility and plans to synthesise data and dissemination strategies. They valued the potential impact of this review on NHS plans for mental health post-pandemic. It has been agreed that the process of this rapid review will be presented to both members for their further comments.

## ETHICS AND DISSEMINATION

As this rapid review will only consider published literature, no ethics approval is needed. Dissemination will be led by the research team and supported by the public member and the wider project advisory group. Results of this review will contribute to reports which will be produced and shared with the National Institute for Health Research (NIHR)Three Research Schools and NIHR Applied Research Collaboration (ARC) North East and North Cumbria (NENC). The findings will be published in peer-reviewed journals and a plain study summary will be disseminated to people receiving mental health care, groups and forum that the project public members are connected, practitioners and commissioners. An abstract will be prepared for academic conferences such as the Society for Academic Primary Care Annual Conference.

## CONCLUSION

This rapid review protocol proposes methods for rapid evidence synthesis. A list of interventions/services/models of care delivered to people with mental health problems during the pandemic will be grouped as “Do”, “Don’t” and “Don’t know” based on the strength of the evidence on effectiveness and acceptability. The proposed rapid review is expected to have an immediate impact on the care of patients by increasing awareness of the impacts associated with service changes and factors enabling high-quality services, in both affected individuals and their service providers. Evidence and practise recommendations produced will be useful in supporting the NHS strategy for planning mental health services in a post-pandemic future, and also in highlighting the needs of service users, potentially the needs of underserved populations. In addition, it will identify important knowledge gaps to help guide research direction.

Accepting the potentially increased risk of biases introduced by rapid review methods is a compromise for many rapid review protocols.^26^ The decision to restrict the evidence review to OECD studies, research published in English and a limited number of databases, and not to undertake a formal quality appraisal exercise will increase the risk of bias and the likelihood that relevant research may be missed or excluded. To help mitigate the limitations of the proposed study design, strategies such as regular team meetings throughout and publishing the protocol in advance of conducting the review have been employed. Furthermore, a quality appraisal may be undertaken retrospectively at a later date for a more comprehensive assessment of the evidence.

## Supporting information

see Appendix

## Data Availability

All data produced in the present study are available upon reasonable request to the authors.

## AUTHORS’ CONTRIBUTIONS

GY and YF conceived the study idea and design. GY drafted the initial manuscript, and all authors reviewed the manuscript and provided input to the final version.

## FUNDING STATEMENT

This project/research/work is supported by the National Institute of Health Research (NIHR) Three Research Schools (award number: MHF018) and the NIHR Applied Research Collaboration (ARC) for the North East and North Cumbria (NENC) (award number N/A). The NIHR is the nation’s largest funder of health and care research and provides the people, facilities and technology that enables research to thrive. <u>Find out more about the NIHR</u>.

## COMPETING INTERESTS STATEMENT

The authors declare that they have no competing interests.

## Notes

### Competing Interest Statement

The authors have declared no competing interest.

### Summary of Updates

1. corresponding author email address was amended. 2. The in-text citation for Appendix was added.

