## Supplementary material for "The impact of changes in mental health services in response to COVID-19 on people with mental health conditions: protocol for a rapid review": see Appendix

**Appendix: Search Strategy**

**PubMed/Medline**

| | **PubMed/Ovid Medline (1950 to date)** |
|---|---|
| **COVID-19** | (2019 nCoV[tiab] OR 2019nCoV[tiab] OR corona virus[tiab] OR corona viruses[tiab] OR coronavirus[tiab] OR coronaviruses[tiab] OR COVID[tiab] OR COVID19[tiab] OR nCov 2019[tiab] OR SARS-CoV2[tiab] OR SARS CoV-2[tiab] OR SARSCoV2[tiab] OR SARSCoV-2[tiab] OR "COVID-19"[Mesh] OR "COVID-19 Testing"[Mesh] OR "COVID-19 Vaccines"[Mesh] OR "Coronavirus"[Mesh:NoExp] OR "Receptors, Coronavirus"[Mesh] OR "SARS-CoV-2"[Mesh] OR "Spike Glycoprotein, Coronavirus"[Mesh]) NOT ("animals"[mh] NOT "humans"[mh]) NOT (editorial[pt] OR newspaper article[pt] |
| **MENTAL HEALTH** | 1. EATING DISORDERS/ or ANOREXIA NERVOSA/ or BINGE-EATING DISORDER/ or BULIMIA NERVOSA/ or FEMALE ATHLETE TRIAD SYNDROME/ or PICA/<br>2. HYPERPHAGIA/ or BULIMIA/<br>3. SELF-INJURIOUS BEHAVIOR/ or SELF MUTILATION/ or SUICIDE/ or SUICIDAL IDEATION/ or SUICIDE, ATTEMPTED/<br>4. MOOD DISORDERS/ or AFFECTIVE DISORDERS, PSYCHOTIC/ or BIPOLAR DISORDER/ or CYCLOTHYMIC DISORDER/ or DEPRESSIVE DISORDER/ or DEPRESSION, POSTPARTUM/ or DEPRESSIVE DISORDER, MAJOR/ or DEPRESSIVE DISORDER, TREATMENT-RESISTANT/ or DYSTHYMIC DISORDER/ or SEASONAL AFFECTIVE DISORDER/<br>5. NEUROTIC DISORDERS/<br>6. DEPRESSION/<br>7. ADJUSTMENT DISORDERS/<br>8. exp ANTIDEPRESSIVE AGENTS/<br>9. ANXIETY DISORDERS/ or AGORAPHOBIA/ or NEUROCIRCULATORY ASTHENIA/ or OBSESSIVE-COMPULSIVE DISORDER/ or OBSESSIVE HOARDING/ or PANIC DISORDER/ or PHOBIC DISORDERS/ or STRESS DISORDERS, TRAUMATIC/ or COMBAT DISORDERS/ or STRESS DISORDERS, POST-TRAUMATIC/ or STRESS DISORDERS, TRAUMATIC, ACUTE/<br>10. ANXIETY/ or ANXIETY, CASTRATION/ or KORO/<br>11. ANXIETY, SEPARATION/<br>12. PANIC/<br>13. exp ANTI-ANXIETY AGENTS/<br>14. SOMATOFORM DISORDERS/ or BODY DYSMORPHIC DISORDERS/ or CONVERSION DISORDER/ or HYPOCHONDRIASIS/ or NEURASTHENIA/<br>15. HYSTERIA/<br>16. MUNCHAUSEN SYNDROME BY PROXY/ or MUNCHAUSEN SYNDROME/<br>17. FATIGUE SYNDROME, CHRONIC/<br>18. OBSESSIVE BEHAVIOR/<br>19. COMPULSIVE BEHAVIOR/ or BEHAVIOR, ADDICTIVE/ |

| | |
|---|---|
| | 20. IMPULSE CONTROL DISORDERS/ or FIRESETTING BEHAVIOR/ or GAMBLING/ or TRICHOTILLOMANIA/<br>21. STRESS, PSYCHOLOGICAL/ or BURNOUT, PROFESSIONAL/<br>22. SEXUAL DYSFUNCTIONS, PSYCHOLOGICAL/ or VAGINISMUS/<br>23. ANHEDONIA/<br>24. AFFECTIVE SYMPTOMS/<br>25. *MENTAL DISORDERS/<br>26. (eating disorder* or anorexia nervosa or bulimi* or binge eat* or (self adj (injur* or mutilat*)) or suicide* or suicidal or parasuicid* or mood disorder* or affective disorder* or bipolar i or bipolar ii or (bipolar and (affective or disorder*)) or mania or manic or cyclothymic* or depression or depressive or dysthymi* or neurotic or neurosis or adjustment disorder* or antidepress* or anxiety disorder* or agoraphobia or obsess* or compulsi* or panic or phobi* or ptsd or posttrauma* or post trauma* or combat or somatoform or somati#ation or medical* unexplained or body dysmorphi* or conversion disorder or hypochondria* or neurastheni* or hysteria or munchausen or chronic fatigue* or gambling or trichotillomania or vaginismus or anhedoni* or affective symptoms or mental disorder* or mental health).ti.<br>27. or/1-26 |
| **LOW AND MIDDLE INCOME COUNTRIES** | (afghanistan OR albania OR algeria OR american samoa OR angola OR "antigua and barbuda" OR antigua OR barbuda OR argentina OR armenia OR armenian OR aruba OR azerbaijan OR bahrain OR bangladesh OR barbados OR republic of belarus OR belarus OR byelarus OR belORussia OR byelORussian OR belize OR british honduras OR benin OR dahomey OR bhutan OR bolivia OR "bosnia and herzegovina" OR bosnia OR herzegovina OR botswana OR bechuanaland OR brazil OR brasil OR bulgaria OR burkina faso OR burkina fasso OR upper volta OR burundi OR urundi OR cabo verde OR cape verde OR cambodia OR kampuchea OR khmer republic OR cameroon OR cameron OR cameroun OR central african republic OR ubangi shari OR chad OR chile OR china OR colombia OR comORos OR comORo islands OR iles comORes OR mayotte OR democratic republic of the congo OR democratic republic congo OR congo OR zaire OR costa rica OR "cote d'ivoire" OR "cote d' ivoire" OR cote divoire OR cote d ivoire OR ivORy coast OR croatia OR cuba OR cyprus OR czech republic OR czechoslovakia OR djibouti OR french somaliland OR dominica OR dominican republic OR ecuadOR OR egypt OR united arab republic OR el salvadOR OR equatORial guinea OR spanish guinea OR eritrea OR estonia OR eswatini OR swaziland OR ethiopia OR fiji OR gabon OR gabonese republic OR gambia OR "geORgia (republic)" OR geORgian OR ghana OR gold coast OR gibraltar OR greece OR grenada OR guam OR guatemala OR guinea OR guinea bissau OR guyana OR british guiana OR haiti OR hispaniola OR honduras OR hungary OR india OR indonesia OR timOR OR iran OR iraq OR isle of man OR jamaica OR jORdan OR kazakhstan OR kazakh OR kenya OR "democratic people's republic of kORea" OR republic of kORea OR nORth kORea OR south kORea OR kORea OR kosovo OR kyrgyzstan OR kirghizia OR kirgizstan OR kyrgyz republic OR kirghiz OR laos OR lao pdr |

| | |
|---|---|
| | OR "lao people's democratic republic" OR latvia OR lebanon OR lebanese republic OR lesotho OR basutoland OR liberia OR libya OR libyan arab jamahiriya OR lithuania OR macau OR macao OR republic of nORth macedonia OR macedonia OR madagascar OR malagasy republic OR malawi OR nyasaland OR malaysia OR malay federation OR malaya federation OR maldives OR indian ocean islands OR indian ocean OR mali OR malta OR micronesia OR federated states of micronesia OR kiribati OR marshall islands OR nauru OR nORthern mariana islands OR palau OR tuvalu OR mauritania OR mauritius OR mexico OR moldova OR moldovian OR mongolia OR montenegro OR mORocco OR ifni OR mozambique OR pORtuguese east africa OR myanmar OR burma OR namibia OR nepal OR netherlands antilles OR nicaragua OR niger OR nigeria OR oman OR muscat OR pakistan OR panama OR papua new guinea OR new guinea OR paraguay OR peru OR philippines OR philipines OR phillipines OR phillippines OR poland OR "polish people's republic" OR pORtugal OR pORtuguese republic OR puerto rico OR romania OR russia OR russian federation OR ussr OR soviet union OR union of soviet socialist republics OR rwanda OR ruanda OR samoa OR pacific islands OR polynesia OR samoan islands OR navigatOR island OR navigatOR islands OR "sao tome and principe" OR saudi arabia OR senegal OR serbia OR seychelles OR sierra leone OR slovakia OR slovak republic OR slovenia OR melanesia OR solomon island OR solomon islands OR nORfolk island OR nORfolk islands OR somalia OR south africa OR south sudan OR sri lanka OR ceylon OR "saint kitts and nevis" OR "st. kitts and nevis" OR saint lucia OR "st. lucia" OR "saint vincent and the grenadines" OR saint vincent OR "st. vincent" OR grenadines OR sudan OR suriname OR surinam OR dutch guiana OR netherlands guiana OR syria OR syrian arab republic OR tajikistan OR tadjikistan OR tadzhikistan OR tadzhik OR tanzania OR tanganyika OR thailand OR siam OR timOR leste OR east timOR OR togo OR togolese republic OR tonga OR "trinidad and tobago" OR trinidad OR tobago OR tunisia OR turkey OR turkmenistan OR turkmen OR uganda OR ukraine OR uruguay OR uzbekistan OR uzbek OR vanuatu OR new hebrides OR venezuela OR vietnam OR viet nam OR middle east OR west bank OR gaza OR palestine OR yemen OR yugoslavia OR zambia OR zimbabwe OR nORthern rhodesia OR global south OR africa south of the sahara OR sub-saharan africa OR subsaharan africa OR africa, central OR central africa OR africa, nORthern OR nORth africa OR nORthern africa OR magreb OR maghrib OR sahara OR africa, southern OR southern africa OR africa, eastern OR east africa OR eastern africa OR africa, western OR west africa OR western africa OR west indies OR indian ocean islands OR caribbean OR central america OR latin america OR "south and central america" OR south america OR asia, central OR central asia OR asia, nORthern OR nORth asia OR nORthern asia OR asia, southeastern OR southeastern asia OR south eastern asia OR southeast asia OR south east asia OR asia, western OR western asia OR europe, eastern OR east europe OR eastern europe OR developing country OR developing countries OR developing nation? OR developing population? OR developing wORld OR less developed countr* OR less developed nation? OR less developed population? OR less developed wORld OR |

| | |
|---|---|
| | lesser developed countr* OR lesser developed nation? OR lesser developed population? OR lesser developed wORld OR under developed countr* OR under developed nation? OR under developed population? OR under developed wORld OR underdeveloped countr* OR underdeveloped nation? OR underdeveloped population? OR underdeveloped wORld OR middle income countr* OR middle income nation? OR middle income population? OR low income countr* OR low income nation? OR low income population? OR lower income countr* OR lower income nation? OR lower income population? OR underserved countr* OR underserved nation? OR underserved population? OR underserved wORld OR under served countr* OR under served nation? OR under served population? OR under served wORld OR deprived countr* OR deprived nation? OR deprived population? OR deprived wORld OR poOR countr* OR poOR nation? OR poOR population? OR poOR wORld OR poORer countr* OR poORer nation? OR poORer population? OR poORer wORld OR developing econom* OR less developed econom* OR lesser developed econom* OR under developed econom* OR underdeveloped econom* OR middle income econom* OR low income econom* OR lower income econom* OR low gdp OR low gnp OR low gross domestic OR low gross national OR lower gdp OR lower gnp OR lower gross domestic OR lower gross national OR lmic OR lmics OR third wORld OR lami countr* OR transitional countr* OR emerging economies OR emerging nation?).ti,ab,sh,kf. |
| **Notes** | [pt=publication type; ab=abstract; ti=title; fs=floating subheading; sh=subject heading; mp=title, original title, abstract, name of substance word, subject heading word, unique identifier] |
